# A normative model representing autistic individuals amidst Autism Spectrum Disorders phenotypic heterogeneity

**DOI:** 10.1101/2021.04.22.21255267

**Authors:** Joana Portolese, Catarina S. Gomes, Vinicius Daguano Gastaldi, Cristiane Silvestre Paula, Sheila C. Caetano, Daniela Bordini, Décio Brunoni, Jair de Jesus Mari, Ricardo Z. N. Vêncio, Helena Brentani

**Affiliations:** LIM23 (Medical Investigation Laboratory 23), University of Sao Paulo Medical School (USP), São Paulo, SP, Brazil; Institute of Psychiatry-University of Sao Paulo, Medical School (FMUSP), São Paulo, SP, Brazil; Development Disorders Program, Universidade Presbiteriana Mackenzie, São Paulo, Brazil; Social Cognition Clinic - TEAMM, Department of Psychiatry, Universidade Federal de São Paulo (UNIFESP), São Paulo, Brazil; Department of Psychiatry, Universidade Federal de São Paulo (UNIFESP), São Paulo, Brazil; Department of Computing and Mathematics FFCLRP, Ribeirão Preto, Universidade de São Paulo (USP), Brazil

**Keywords:** Autism Spectrum Disorder, Phenotypic Heterogeneity, Normative modeling, Principal Component Analysis

## Abstract

Approaches to deal and understand Autism Spectrum Disorder (ASD) phenotypic heterogeneity, quantitatively and multidimensionally, are in need. Being able to access a specific individual relative to a normative reference ASD sample would provide a severity estimate that takes into account the spectrum variance. We propose such an approach analyzing the principal components of variance observable in a clinical reference sample. Using phenotypic data available in a comprehensive reference sample, the Simons Simplex Collection (*n*=2744 individuals), we performed Principal Component Analysis (PCA). The PCA considered ASD core-symptoms (accessed by ADI-R), important clinical features (accessed by VABS and CBCL) and IQ. PCA-projected dimensions supported a normative modeling where a multivariate normal distribution was used to calculate percentiles. An additional phenotypically homogeneous sample (ASD, IQ<75, 6-7yr, *n*=60) is presented as a case study to illustrate the phenotypic heterogeneity assessment and individual placement under the normative modeling approach. Three PCs embedded 72% of the normative sample variance, interpreted based on correlations (>0.50) with clinical features as: Social Functionality (39%), Behavioral Disturbance (18%) and Communication Problems (15%). A Multidimensional Severity Score (MSS) to evaluate new prospective single subjects was developed based on percentiles. Additionally, the disequilibrium among PCA-projected dimensions gave rise to an individualized Imbalance Score (ImS). The approach, named TEAplot, is implemented in user-friendly free software and was illustrated in a homogenous independent sample. Our approach proposes a basis for patient monitoring in clinical practice, guides research sample selection and pushes the field towards personalized precision medicine.

**Lay Summary:** Most families or clinicians already heard the now adage: “If you’ve met one person with autism, you’ve met one person with autism”. The phenotypic heterogeneity presented by the Autism Spectrum Disorders (ASD) is a challenge to research and clinical practice. Here in this work we summon established mathematical tools from the Machine Learning field to help one to organize the principal components of such variability. These mathematical tools were applied to a comprehensive database of autistic individuals’ mensurable profiles (cognitive, emotional, behavioural, and so on) maintained by the Simons Foundation Autism Research Initiative (SFARI). Using this normative model one can quantitatively estimate how a given individual person fits into the whole, as pediatricians often do by evaluating growth charts, a tool we named TEAplot. We made freely available Excel/Libreoffice spreadsheets that calculate our proposed Multidimensional Severity Score in order to effectively engage the research and clinical communities. The TEAplot model is a step towards a personalized precision medicine approach for ASD.

## INTRODUCTION

Autism Spectrum Disorder (ASD) is a multifactorial disorder, with a complex genetic architecture and great variability in clinical presentation (Masi et al., 2017). This great heterogeneity brings difficulties for diagnosis, prognosis, and therapeutic response both in clinical practice and in the research area (Havdahl et al., 2016; Amaral et al., 2019). In the Diagnostic and Statistical Manual of Mental Disorders, 5th edition (American Psychiatric Association, 2013), ASD is classified as a spectrum of changes in two dimensions: deficiencies in social communication and restricted and repetitive interests or behaviors. Different studies contributed to the DSM-5 idea of a spectrum with a severity gradient model (Hus et al., 2007; Lord & Jones, 2012; Bellinger & Smith, 2001; Constantino et al., 2004; Hus & Lord, 2013). The important role of other than ASD core symptoms, and their impact on the severity of ASD clinical presentation, has been recognized in DSM-5. They are represented as modifiers such as IQ and language capacity (Lord et al., 2004; Grzadzinski et al., 2013). The Childhood Autism Rating Scales (CARS) (Schopler et al., 1994) yielded an overall ASD severity score and, the total scores in the Autism Diagnostic Observation Schedule (ADOS) were revised to provide a continuous measure of severity less influenced by child characteristics such as age and language yielding the ADOS-2 Calibrated Severity Scores (CSS) (Gotham et al., 2009; Hus et al., 2014).

The various levels of functioning have also been recognized as important factors, contributing to severity and ASD trajectories (Bitsika et al., 2008; Kanne et al., 2011; Farmer et al., 2018). Another contributing characteristic is the presence of emotional/behavioral problems (Georgiades et al., 2011). Disruptive behaviors in children with ASD are relatively common, being presented by a quarter to a third of them, including outbursts of anger, irritability, opposing behavior, and aggression (White et al., 2014). These behaviors can affect scores in diagnostic algorithms (Hus et al., 2013) and, in conjunction with other clinical data, contribute to characterize ASD clinical heterogeneity (Waddington et al., 2018; Lombardo et al., 2019). Different studies have been using IQ, functionality, emotional/behavioral problems, and other phenotypic measures along with core symptoms to find subgroups and better describe ASD clinical heterogeneity, characterizing the severity spectrum (Syriopoulou-Delli & Papaefstathiou, 2020; Al-Jabery et al., 2016; Stevens et al., 2019). However, it is important to note that these studies do not focus on an individual patient.

Recently, the promising approach of “normative modeling” has been put forward to characterize the heterogeneity of psychiatric disorders (Marquand et al., 2019). This approach uses a probabilistic model to determine a distribution/variation of some symptoms and biological characteristics given relevant covariables. Using normative representations, the degree to which individuals deviate from reference population ranges can be assessed in a personalized fashion.

The studies that carried out the normative modeling approach to better characterize psychiatric disorders used healthy individuals to create normative population ranges (reviewed in [Marquand et al., 2019]). However, it is also possible to create a map of normative ranges to understand the heterogeneity of a specific clinical presentation, *e*.*g*. in ASD, in contrast to a neurotypical population. In this context, a patient can be evaluated on the normative map, giving clinicians an idea of relative positions, and orienting not only the comparison among individuals but also individual changes over time.

In this work, we propose a two-step integrated approach to (i) combine measures of phenotype variability that contribute to clinical heterogeneity in ASD and (ii) use it with normative modeling to assess severity. Considering severity as a multidimensional construct, we used Principal Component Analysis on measures of clinical heterogeneity impacting severity. A large publicly available clinical reference sample from Simons Foundation Autism Research Initiative was used as the normative database. Based on that we propose severity and imbalance scores that take into account ASD phenotypic heterogeneity. Finally, we demonstrate the proposed approach usage in a case study using an ASD homogeneous sub-sample of a previously published study (Bordini et al., 2020).

## MATERIAL AND METHODS

### Participants

We used 2744 (out of 2857, 86% male) probands with no missing data from the Simons Simplex Collection (SSC) sample (version 15.0) from the Simons Foundation Autism Research Initiative (SFARI) database as a clinical reference sample (Fischbach & Lord, 2010). In order to build a case study, we use the baseline data from a recently published study aiming for as much homogeneity as possible: 60 patients (out of 66), 6 to 7 years old, ASD diagnosis, intellectual disability (IQ<75) and low functionality (Bordini et al., 2020). Table S1 and Table S9 describe scales/questionnaires mean and standard deviation scores, relative to the reference and case samples, respectively.

### Assessment tools

In order to measure ASD symptoms, functionality, and the behavioral profile we used Autism Diagnostic Interview-Revised (ADI-R) (Lord et al., 1994), Vineland Adaptive Behavior Scales (VABS) (Sparrow et al., 1984), and the Child Behavior Checklist (CBCL) (Achenbach & Rescorla, 2001). Standardized scores of VABS and CBCL were used. Cognitive assessment was estimated by the Total Intelligence Quotient (IQ). Concerning the SSC database, Full-Scale Deviation IQ data was used whenever available, and alternatively, Full-Scale Ratio IQ was used instead if not. Concerning the Brazilian case study sample, a standardized, age-corrected and Portuguese-validated tool was used (Bordini et al., 2020). Details on IQ instruments used are available in Supplementary Methods.

### Principal Component Analysis

In order to capture the most prominent sources of variability in the normative reference dataset a Principal Component Analysis (PCA) was performed. The 9 input variables were: Socialization, Communication, and Daily Living Skills (VABS domains); Socialization, Communication, and Restricted and Repetitive Behaviors (ADI-R domains); total IQ and the t-score values of CBCL Internalizing and Externalizing problems. The Principal Components (PC) that together explained up to 70% of the sample’s variability were selected. In order to assign interpretability to each PC, we consider the original variables presenting absolute correlations with it greater than 0.5 as relevant. The information from the PCA performed in the clinical reference sample was used to project new observations/patients into the PC-rotated coordinate system. The final mean-centered variance-standardized lower dimensional map, core of our approach, was named TEAplot.

### Normative model

Gaussian Modeling was used to derive a normative model that captures reference sample’s phenotypic variation by fitting a multivariate normal density to the PCA derived coordinates. A special direction on the lower dimensional map, from all negative value corner to all positive corner, was defined as a natural gradient direction of clinical severity presentation. The probability of being worst in this direction is calculated for any individual using a simple univariate normal density since, along this diagonal direction, the lower dimensional multivariate normal density is spherically symmetrical and reduces itself to the univariate case. The probability quantile calculated along the aforementioned “special” axis defines a relative severity score (details available as Supplementary Material).

### Multidimensional Severity Score (MSS) and Imbalance Score (ImS)

In the TEAplot, each patient is represented by a point at the lower dimensional coordinate system of Z-score transformed PCs. A directed diagonal axis, which reflects general phenotypic severity, was defined from the all negative octant to the all positive octant. In order to estimate the degree of phenotypic severity, we calculate a 1-dimensional projection of a patient point into the aforementioned axis. This projected point is the input variable for an accumulated probability integration based on the Gaussian modeling. The integrated quantile is then multiplied by 10 to yield the proposed Multidimensional Severity Score (MSS). In order to quantify the imbalance among the lower dimensional coordinates in TEAplot, we propose a score based on entropy (base chosen to normalize results in [0,1]) multiplied by 10, named Imbalance Score (ImS). In summary, each patient received 5 values: scores on three principal components normalized to Z-Scores, a score measuring the disequilibrium among the aforementioned three (ImS), and a score which represents the overall severity when compared to the reference sample (MSS). Expanded derivations and details on calculating the scores can be found in Supplementary Material.

### Statistical and Computational analysis

Multivariate statistical methods and graphics were implemented using R statistical language (v4.0.3) scripts. The PCA implementation used was `prcomp` with all default parameters except scaling, set to `TRUE`. Excel/LibreOffice. `xls` files are freely available as Supplemental Material to allow one to take advantage of the proposed normative approach for individual patients.

## RESULTS

In the following we will describe results obtained by the proposed approach. Figure 1 shows an overview of the 3 main phases of our approach: PCA application on SSC sample, normative modeling and personalized evaluation strategy.

**Figure 1:**
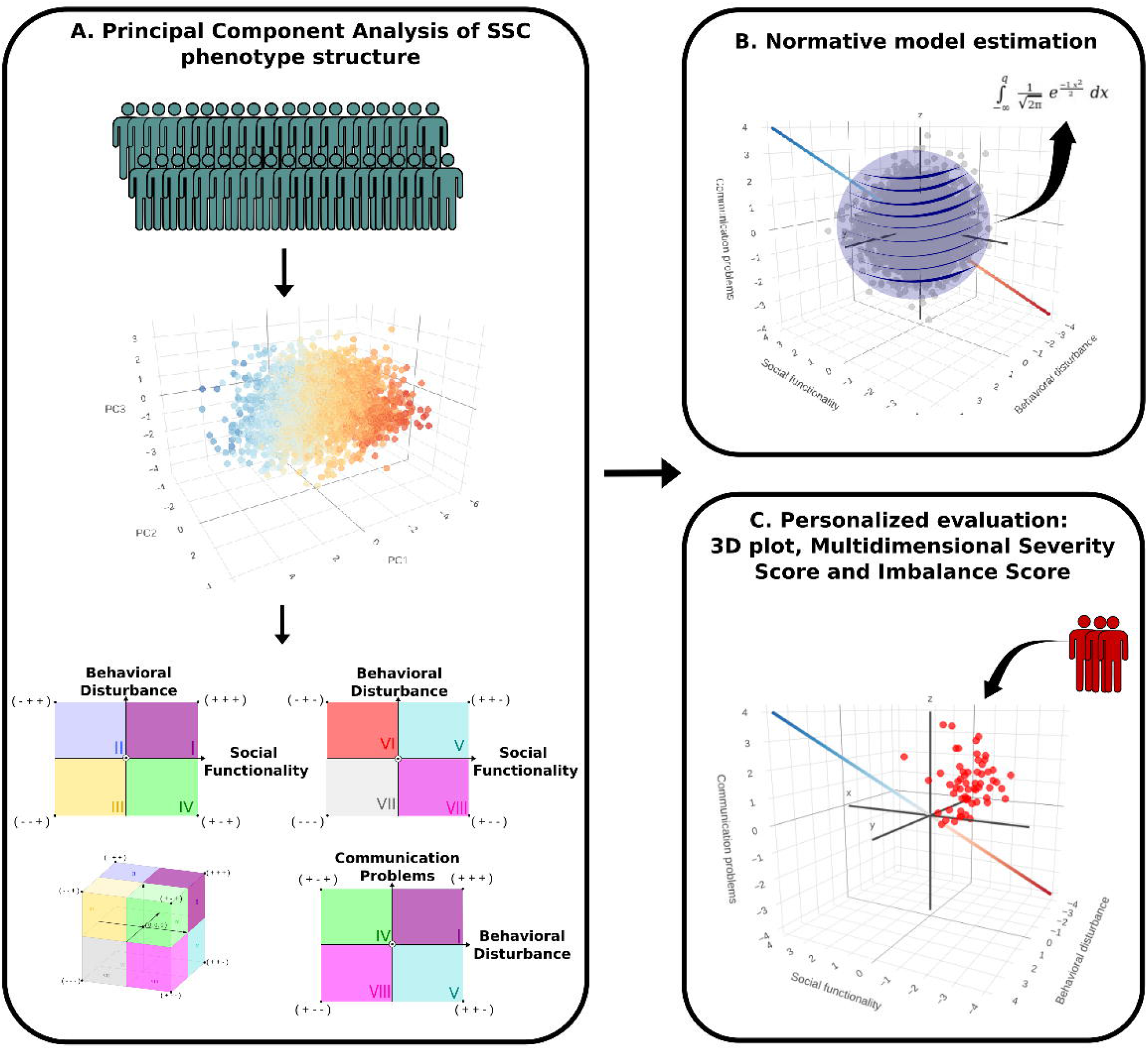
Overview of the method. Panel A shows the phenotypic heterogeneity map construction, based on the SSC patients coordinates in three principal components. Each octant of the 3D PC coordinate system has an clinical interpretation, resulting in three axes of phenotypic variability: “Social Functionality”, “Behavioral Disturbance”, and “Communication Problems”. Gaussian Modeling was used to derive a normative model that captures reference sample’s phenotypic variation by fitting a multivariate normal density to the PCA derived coordinates, concerning a special direction on the 3-dimensional map defined as a natural gradient direction of clinical severity presentation (panel B). Any new patient can be mapped in the 3D space endowed with natural clinical interpretation and receive a “Multidimensional Severity Score”, and a score which reflects the existent relationship between the clinical aspects reflected by each PC: the “Imbalance Score” (panel C).

### Principal Component Analysis

In order to get a multidimensional view, a PCA using the subscales measures of the selected phenotypic instruments as input was performed and the first three principal components, that together explained around 70% of the SSC sample variability, were selected (Table 1 and Table S2). Loading contribution of each Principal Component (PC) showed that: (i) PC1 has a greater correlation (>0.65) with VABS subscores, total IQ, and ADI-R Socialization; (ii) PC2 has a greater correlation (>0.50) with CBCL Internalizing and Externalizing Problems and with ADI-R Restricted and Repetitive Behaviors; and **(iii)** PC3 has a greater correlation (>0.70) with ADI-R Communication. Aiming to test the PCA based phenotypic structure stability, the same analysis was performed using a random SSC sample subset. When using from 30% up to 80% of the SSC sample, all PCA results remain essentially the same (Table S3 - Table S8).

**Table 1:** Principal Component Analysis (PCA) of SSC reference sample. The first three principal components, which sum up to approximately 70% of the sample variability, were considered by the model. Pearson correlations between components and original input variables are shown, with >0.50 in bold. An expanded version with all nine components is available in Supplementary TableS2.

| Component | 1 | 2 | 3 |
| --- | --- | --- | --- |
| *Eigenvalue* | 3.49 | 1.65 | 1.39 |
| *Explained Variance (%)* | 39 | 18 | 15 |
| *Cumulative Explained Variance (%)* | 39 | 57 | 72 |
| Correlations |  |  |  |
| *VABS Communication* | **0.89** | -0.16 | 0.17 |
| *VABS Socialization* | **0.87** | 0.04 | 0.12 |
| *VABS Daily Living Skills* | **0.85** | -0.13 | 0.19 |
| *Total IQ* | **0.78** | -0.29 | 0.12 |
| *ADI-R Socialization* | **-0.69** | -0.18 | 0.43 |
| *ADI-R Communication* | -0.31 | -0.34 | **0.73** |
| *ADI-R Restricted and Repetitive Behavior* | -0.17 | **-0.52** | 0.45 |
| *CBCL Internalizing Problems* | -0.04 | **-0.78** | -0.39 |
| *CBCL Externalizing Problems* | -0.14 | **-0.71** | -0.48 |

Note: VABS = Vineland Adaptive Behavior Scales; ADI-R = Autism Diagnostic Interview-Revised; IQ = Intelligence Quotient; CBCL = Child Behavior Checklist.

Although PCs are linear combinations of original variables with no immediate pairing with instruments, the examination of individual top correlated variables with PCs yield some insights. Figure S1 to Figure S9 show SSC patients in their (PC1, PC2, PC3) tridimensional coordinates (TEAplot) color-coded according to increasing original variable values. Individuals located in the negative score octants on PC1 have worse scores on the subscales of the instruments that reflect aspects of social functionality such as VABS, Total IQ, and ADI-R Socialization (Figure S1 - Figure S5). Individuals located in the negative score octants on PC2 have worse scores on CBCL internalizing and externalizing problems, which reflect aspects of emotional and behavioral symptoms, and on ADI-R Restricted and Repetitive Behaviors (Figure S6 - Figure S8). Individuals located in the negative score octants in PC3 have worse scores in the ADI-R Communication (Figure S9).

Therefore, by sectorizing the 3-dimensional projection of a PC coordinate system into eight octants, the association between the phenotypic presentation and PC values can be generalized as shown in Figure 2.

**Figure 2:**
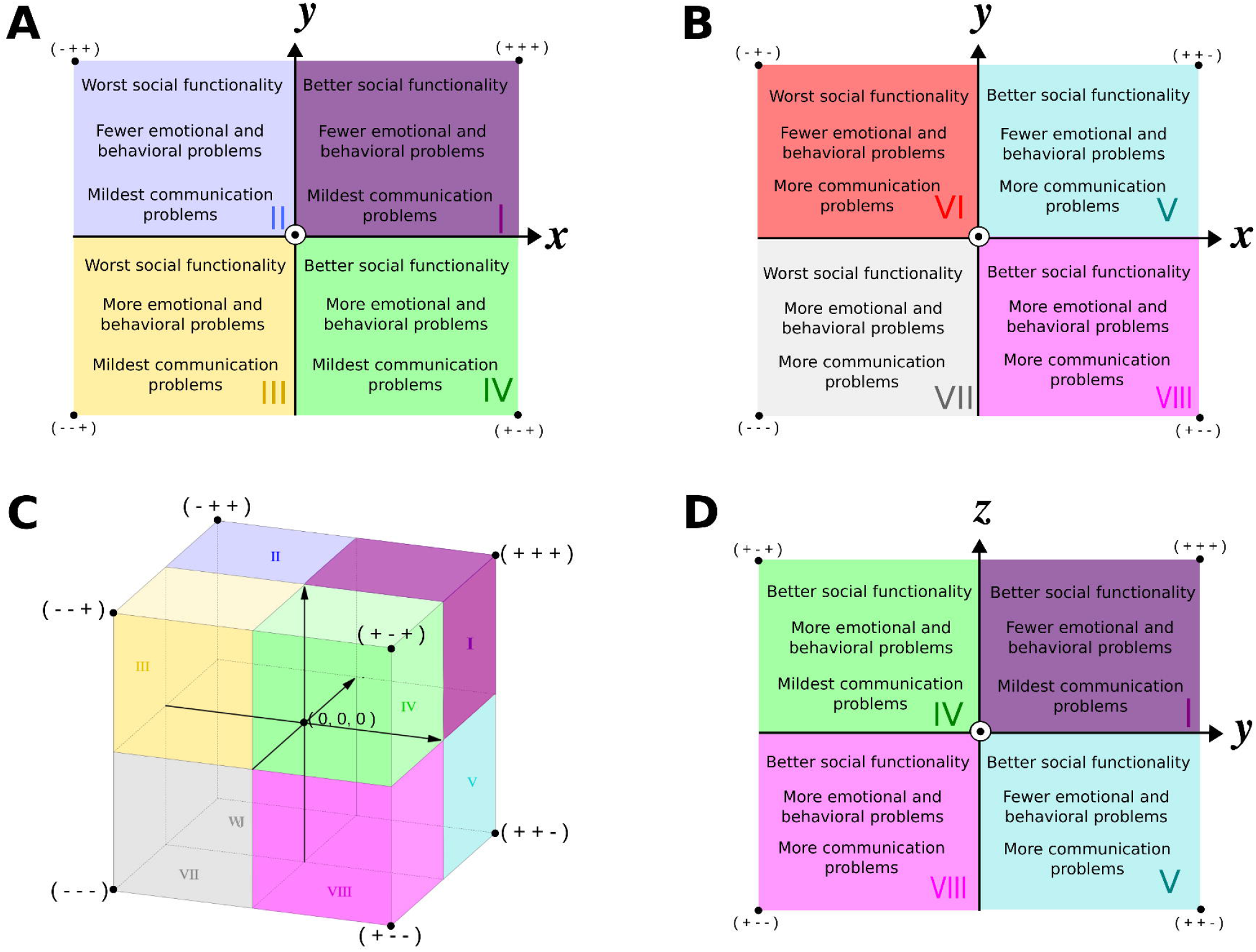
Clinical and conceptual implications of dimensional representation of autistic individuals. The tridimensional space proposed to holistically represent individuals is divided into 8 octants, labeled from I to VIII (panel C). Schematically, x-, y-, and z-axis embed principal components 1, 2, and 3, respectively. Two-dimensional views of the space are shown for clarity (panels A, B and D) along with octant clinical interpretation (text inside). Each octant corner indicates with “+” or “-” signals qualitative better or worse clinical status for the 3 dimensions. The circled dot at the origin represents an axis directed towards outside the plane shown and z-axis positive and negative octants (panel A and B, respectively) are shown separately for clarity. Adapted from Wikipedia’s ‘octant’ entry.

PC1 represents the variation from individuals with less social functionality to the more functional ones and it can be interpreted as **“Social Functionality”**. In the PC2 axis, we observe a gradation of individuals with more internalizing or externalizing behavior problems and stereotyping behaviors to individuals with fewer problems, that is, better adequacy of emotional and behavioral regulation. Thus, PC2 can be interpreted as **“Behavioral Disturbance”**. Finally, the PC3 axis was sign-adjusted so there is a gradation from individuals with more communication problems to individuals with fewer communication problems, assessed by the ADI-R sub-item. Therefore, the PC3 can be interpreted as **“Communication Problems”**.

### Quantifying severity and phenotype heterogeneity using a Multidimensional Severity Score and an Imbalance Score

A general direction, from worst to best general clinical presentations, which goes from octant VII’s vertice (all negative components) to octant I’s vertice (all positive components) was observed (Figure S10).

Along with patient visualization in a map endowed with natural clinical interpretation, this representation also allows a normative modeling. The phenotypic heterogeneity embedded in the clinical reference sample was modeled by a probability density function and its quantiles provide a simple way to assess prospective patients’ severity of clinical presentation. The modeled fraction of “worst of” patients multiplied by 10 returns a score for a given prospective individual that we named “**Multidimensional Severity Score” (MSS)**. MSS is a relative severity gradient from 0 to 10. In contrast to standard quantiles used in growth charts, which are two-sided, the proposed MSS score is one-sided since it assumes the all negative and all positive octants as the worst and best possible clinical presentations, respectively, increasing along this direction (Figure 2 and Figures S10-S19). The distribution of MSS calculated for the SSC subjects is essentially uniform (Figure S20) meaning that the proposed score increases approximately linearly along the diagonal severity axis.

In spite of ADOS being developed as a diagnosis algorithm, the ADOS-2 standardized Calibrated Severity Scores (CSS) (Gotham et al., 2009; Hus et al., 2014) has been modified to assess a patient’s autistic symptomatology severity. Our comprehensive MSS score shows no direct/simple relationship with CSS score in the large SSC sample (Figure S21).

We also defined a score which reflects the existent relationship between the clinical aspects reflected by each PC (Social Functionality, Behavioral Disturbance, and Communication Problems). Regardless of the severity level estimated, a patient’s deficit could be concentrated in a single component. In order to capture this imbalance we used simple entropy measurement considering all 3 components. Therefore, salient disparities among components can be quantified as a 0 to 10 score named “**Imbalance Score**” **(ImS)**. Patients with ImS near 10 have a more equilibrated relationship among the three PC axes. Conversely, patients with ImS near 0 hold more information in one specific PC axis so there is a disequilibrium among the three variability axes.

### Case presentation

The TEAplot 3-dimensional space was obtained using the SSC normative sample but the main advantage of our approach resides in its translational power to any similarly obtained ASD patient. Our local sample has individuals accessed with equivalent standardized instruments therefore can be mapped in the PCA-derived normative space using the same rotation eigenvalue built rotation matrix and normalization coefficients. Therefore, we assigned our 60 patients on the defined phenotypic heterogeneity map (Figure 3).

**Figure 3:**
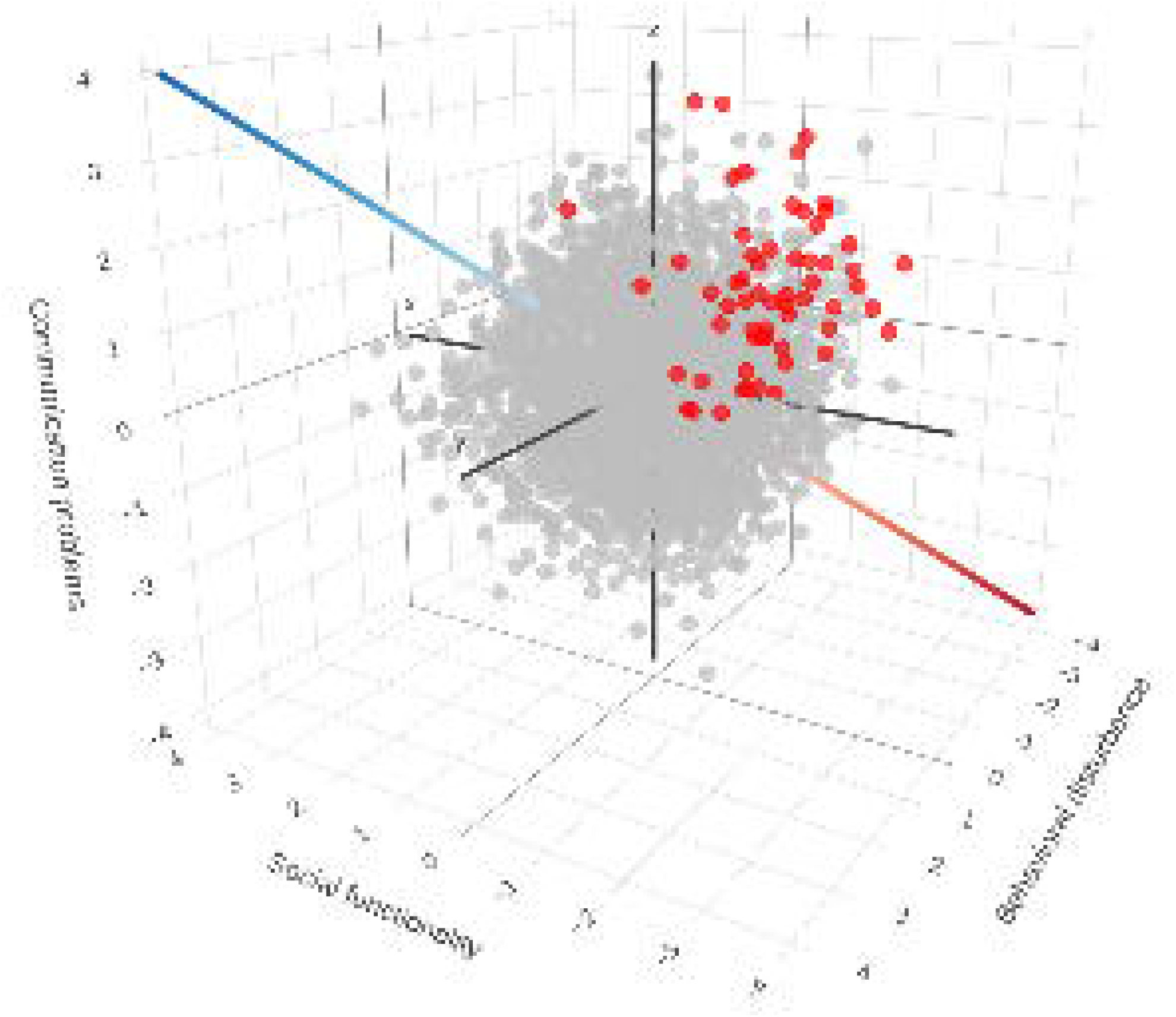
Visualization of case presentation patients on the map of phenotypic heterogeneity under normative modeling. The case presentation patients are shown in red. The principal components account for 73% of total variance distributed as: Social Functionality (39%), Behavioral Disturbance (18%) and Communication Problems (15%).

Compared to the reference map, most patients are in the octant that reflects worst clinical presentations concerning Social Functionality and Behavioral Disturbance but with less severe Communication Problems. It is notable that although these patients would be clinically considered as homogeneous, there is wide variability considering the three TEAplot axes. Besides visualizing the patients on the phenotypic heterogeneity map and empirically analyze the relationships between their position and the patient’s clinical presentation, we quantified the deviations from phenotypic heterogeneity concerning the reference clinical sample using the MSS and the existent heterogeneity in the three axes of variation using the ImS (presented in detail in Table S10).

In order to clarify how score interpretation reflects clinical observations, we highlight some patients (Table 2). Patients ID-07 and ID-46 are quantitatively different regarding their severity placement in the spectrum: MSS 2.1 and 9.5, respectively. ID-07 has more negative values in the first 2 PCs and with less imbalance (9.7), whereas ID-46 has PC1 and PC2 negative, but more positive results from PC3, with greater imbalance (4.9). The discrepancies between the clinical scoring range in daily living skills, in the IQ level, and in externalizing problems, account for the lower imbalance of patient ID-07 compared with ID-46. Another example are ID-77 and ID-78 that have a total ADI-R score of 51 and 54, respectively. Both patients have a similar degree of core symptoms. In order to better understand possible differences between them we could look for scales associated with the clinical severity profiles: IQ, sub-items of VABS, and CBCL. Comparing the results of these patients, we observe that ID-78 has lower IQ, more repetitive and stereotyped behaviors, and more internalizing and externalizing behaviors, with behavioral problems in the CBCL clinical range, when compared to the ID-77, and probably this patient would be allocated in the severity spectrum as a worst presentation. Using the proposed normative model, one is able to quantify the difference since ID-77 and ID-78 have MSS of 3.8 and 2.1, respectively. Meanwhile, heterogeneity differences observed in the clinical assessment can be compared using their PC values in TEAplot space.

**Table 2:** Phenotypic deviation index for some patients of the case presentation sample. *Information not used as input in the model.

| IDs | VABS<br>Com | VABS<br>Soc | VABS<br>DLS | Total<br>ADI-R* | ADI-R<br>Soc | ADI-R<br>Com | ADI-R<br>RRB | Total<br>IQ | CBCL<br>Int | CBCL<br>Ext | PC1 | PC2 | PC3 | MSS | ImS |
| --- | --- | --- | --- | --- | --- | --- | --- | --- | --- | --- | --- | --- | --- | --- | --- |
| ID 01 | 39 | 48 | 37 | 60 | 30 | 14 | 13 | 49 | 76 | 67 | -2.55 | -1.46 | 0.65 | 0.3 | 8.8 |
| ID 02 | 39 | 32 | 88 | 52 | 27 | 14 | 10 | 50 | 80 | 77 | -1.84 | -1.94 | 1.25 | 0.7 | 9.8 |
| ID 07 | 67 | 59 | 56 | 48 | 24 | 13 | 10 | 55 | 79 | 54 | -1.13 | -0.86 | 0.62 | 2.1 | 9.7 |
| ID 08 | 52 | 54 | 59 | 53 | 25 | 13 | 10 | 67 | 58 | 56 | -1.36 | 0.07 | 0.11 | 2.5 | 4.2 |
| ID 21 | 51 | 55 | 62 | 48 | 24 | 13 | 10 | 55 | 61 | 52 | -1.36 | 0.17 | 0.13 | 2.7 | 5.6 |
| ID 23 | 63 | 68 | 84 | 48 | 24 | 13 | 10 | 51 | 58 | 54 | -0.53 | 0.08 | -0.27 | 3.4 | 8.2 |
| ID 46 | 58 | 80 | 55 | 31 | 20 | 5 | 2 | 69 | 73 | 83 | -0.42 | -0.24 | 3.50 | 9.5 | 4.9 |
| ID 75 | 46 | 52 | 55 | 51 | 28 | 14 | 4 | 62 | 78 | 77 | -1.72 | -1.01 | 2.00 | 3.4 | 9.7 |
| ID 77 | 47 | 51 | 45 | 51 | 24 | 19 | 3 | 70 | 63 | 62 | -1.71 | 0.29 | 0.88 | 3.8 | 8.2 |
| ID 78 | 47 | 57 | 53 | 54 | 23 | 20 | 6 | 59 | 65 | 67 | -1.60 | -0.38 | 0.56 | 2.1 | 8.3 |

Note: VABS Com = Vineland Adaptive Behavior Scales Communication; VABS Soc = Vineland Adaptive Behavior Scales Socialization; VABS DLS = Vineland Adaptive Behavior Scales Daily Living Skills; ADI-R Soc = Autism Diagnostic Interview-Revised Socialization; ADI-R Com = Autism Diagnostic Interview-Revised Communication; ADI-R RRB = Autism Diagnostic Interview-Revised Repetitive Restricted Behavior; IQ = Intelligence Quotient; CBCL Int = Child Behavior Checklist Internalizing Problems; CBCL Ext = Child Behavior Checklist Externalizing Problems; PC = Principal Component; MSS = Multidimensional Severity Score.

Clinically, comparing patient ID-75 with ID-23, the scores of the sub-items of the ADI-R Socialization and Communication are similar, but the ID-23 presents higher scores in the VABS, and results of the CBCL Internalizing and Externalizing problems are in the non-clinical range. However, the ID-75 has higher IQ scores and better results from Restricted Repetitive Behavior. Would two health personnel performing the evaluation allocate them equally in the severity spectrum? The MSS shows that they are at the same point. We can also say that they have some differences associated with discrepancies between the clinical scoring ranges in Daily Living Skills, Internalizing and Externalizing problems and the clinical scoring ranges in the Restricted Repetitive Behavior of the ADI-R and the IQ, once the Imbalance Score of the ID-75 patient is lower than the ID-23. Comparing the results of ID-01 and ID-02, we can say that ID-02 is more functional regarding daily living skills although with more behavioral problems. It would be difficult to compare their allocation in the severity spectrum, however the normative model shows us that they have a similar MSS (ID-01 with 0.3 and ID-02 with 0.7), but ID-01 is not only slightly more severe but also has more imbalance (ImS 8.8 compared with 9.8 of ID-02). As expected, the PCs scores comparison will render the same clinical direction observed with the scales score. Comparing the IQ, VABS, and CBCL results of ID-08 and ID-21, we can say that the ID-08 has a higher IQ, despite lower results in the score of activities of daily living when compared to the ID-21. On the other hand, ID-21 presents slightly more problems in internalizing behaviors, when compared to ID-08. The proposed model quantifies our clinical view. ID-08 and ID-21 patients showed similar severity scores (2.5 and 2.7, respectively), and the ImS of 4.2 for ID-08 and 5.6 for ID-21 showed that both individuals had discrepancies between the components.

The clinical normative map and deviation scores can also inform us about a quantitative patient’s longitudinal follow-up. By allocating a patient, ID-28, diagnosed with ASD and assessed at six years old and reevaluated using the same scales at seven years old, using the MSS and ImS we were able to quantify his trajectory based on the reference clinical sample (Figure S22). At the time of assessment (*t*_1_), the patient had MSS of 8, which evolved to 9.1 one year after (*t*_2_) (Table S11). At the first assessment (*t*_1_), the patient had an imbalance score ImS of 7.0, which went to 9.2 one year after diagnosis (*t*_2_). This is an interesting example where it was possible to observe a small decrease in severity over time and that it was reflected in all PC components, showing a changing trajectory increasing phenotypic relationship balance.

## DISCUSSION

Selecting principal components which explain up to 70% of the variability of the reference sample was used to create a map of clinical heterogeneity components contributing to the severity spectrum. Based on normative modeling using the principal components of phenotypic heterogeneity in ASD, any patient evaluated by the phenotypic scales used can receive a severity score. To our knowledge, this is the first study that proposed a systematic way to combine: (i) quantitative access to the phenotype variability that contribute to the ASD clinical presentation heterogeneity; and (ii) normative modeling methods, thus allowing one to map and quantify an individual’s clinical severity in a defined normative space.

We sought to use PCA to reduce the dimensionality of characteristics recurrently associated with the heterogeneity and severity of ASD clinical presentation in the literature. The first component reflects works in the literature that showed that IQ and adaptive behavior are correlated with socialization scores and interfere with clinical severity (Bitsika et al., 2008; Kanne et al., 2011). Interestingly, VABS’s weight in this component is corroborated by a longitudinal study that has shown an important role of the adaptive functioning in delineating ASD cases with different developmental trajectories (Farmer et al., 2018). Despite the fact that Intellectual Disability (ID) estimates in the ASD population have progressively decreased in recent decades to one-third of individuals with ASD, with 25% and 44% of individuals estimated in the ID/borderline and medium/above-average ranges, respectively (Baio et al., 2018), longitudinal study indicate low IQ as a factor of poor prognosis (Gotham et al., 2012). Nonetheless, a higher IQ can be a necessary but not sufficient condition for positive results in areas of functioning, such as relationships, employment, and independence (Howlin et al., 2013). Tillmann et al., (2019) tested unique predictors of adaptive functioning as measured by the VABS and the discrepancy between IQ and adaptive functioning in ASD, and showed that socio-communicative symptoms, but not sensory/repetitive symptoms or concomitant psychiatric symptoms (anxiety, depression, and ADHD), are associated with less adaptive functioning and greater discrepancies in adaptive capacity.

The second component refers to internalizing and externalizing problems and Restricted Repetitive Behavior (RRB). RRB has been considered a separate dimension from social-communication before and after the DSM-5 classification, accordingly, it is expected that it will be more correlated with a different PC from sociability. Psychiatric and medical comorbidities are present in about 70% of individuals with ASD (Pandolfi et al., 2012), anxiety disorders are the most common, affecting about 40% of children and adolescents with ASD (Gotham et al., 2013). In addition to anxiety, depression is also highly prevalent throughout life, occurring in 17% to 70% (Hollocks et al., 2019). Deficiencies in emotional regulation are a risk factor for anxiety in ASD (Neuhaus et al., 2019). Georgiades et al. (2011) examined the phenotypic overlap between the main diagnostic characteristics and the emotional/behavioral problems in ASD using PCA. As we observed, the PC associated with emotional behavior was uncorrelated to intellectual, functional adaptive, and structural language skills of children. Individuals with ID are at higher risk of presenting behavioral problems, but it has been shown that children affected with ID have a higher risk of presenting only some behavioral problems such as self-injury and abnormal eating behaviors, validating that they represent distinct components (Kurzius-Spencer et al., 2018).

It has been suggested that lack of flexibility and RRB may contribute to emotion dysregulation in children with ASD (Mazefsky et al., 2013). It was also suggested that autistic individuals can sustain hyper-focus on upsetting past events and that a correlation between ASD core symptoms and anger rumination exists (Patel et al., 2017; Ibrahim et al., 2019). In these, and other studies, authors have been discussing how RRBs and behavioral problems interact contributing to ASD trajectories (Pugliese et al., 2016; Bos et al., 2018).

The third and last component was represented by ADI-R Communication. Considering that in DSM-5 some items of communication were included in the dimension of sociability and others in the dimension of repetitive and stereotyped behaviors, it may appear that our findings disagree with the two-dimensional model (Lord & Bishop, 2015). However, we think the opposite. First, the two-dimensional model proposed in DSM-5 is for diagnosis, however, as already discussed, language ability is a modifier since it is not necessary for diagnosis although interferes with severity and heterogeneity (Visser et al., 2017). Our scores are not for diagnosis, but to characterize heterogeneity and quantifies severity, but the fact that PC3 contributes to the first and second component, featuring a specific component, seems fully in accordance with DSM-5. We suggest that what is represented in PC3, that is not represented in the other components, is precisely the language ability part of communication. Previous work seeking to create ASD ontologies mapped 27% of the items in ADI-R, specifically to language ability (McCray et al., 2014). Moreover, ADI-R scoring system has one subdomain of communication coded for children at all language levels: gesture and play. Other subdomains are considered separately for children with phrases (verbal) and children with single words or no words at all (nonverbal) (Lord et al., 1994). For verbal children, besides gestures and play, language ability/skills is also evaluated. In the SSC 88% are verbal and 12% nonverbal, so language ability was evaluated and contributed to finding components in most cases in SSC normative samples. Language abilities are not necessary for the diagnosis, but 30% of ASD children remain minimally verbal when entering school (Norrelgen et al., 2015; Rose et al., 2016). A recent study concluded that lower adaptive results, higher IQ (measured over time), and language ability in childhood, tend to predict autonomy results in adulthood (Simonoff et al., 2020).

Since we are proposing a severity score, although ADOS is a diagnosis instrument, we compare the MSS with the CSS score, a modified ADOS score to assess severity. We didn’t observe any clear relationship between scores. This would be expected considering that ADOS CSS evaluates only ASD core symptoms (Gotham et al., 2009; Hus et al., 2014) and MSS takes into account other non core ASD symptoms. Different studies showed that language ability, IQ and Vineland scores are the ones with greater variability and impact in ASD trajectories and not ASD core symptoms (Lombardo et al., 2019; Syriopoulou-Delli & Papaefstathiou, 2020; Al-Jabery et al., 2016; Stevens et al., 2019).

The study results increase the existing literature on ASD by emphasizing the importance of assessing general emotional/behavioral problems, functionality, IQ and language ability in conjunction with the main diagnostic symptoms in children with ASD to characterize the clinical heterogeneity and assessment of severity. The mapping proposed here allows clinical phenotypes to be dissected along the most relevant axes of variation. Thus, we can map the variability between different domains of operation and compare individuals regarding this variability as proposed by studies of normative models (Marquand et al., 2019).

In this way, we were able to develop a map where patients with ASD can be allocated and their phenotypic relationships evaluated, which allows for the best characterization of an individual patient as well as for making comparisons in a standardized view between cases as presented in the case study, even in a homogeneous clinical sample.

The putative translational success of this normative model is predicated upon the normative sample power to serve as a relevant reference population. The SFARI SSC database is a renowned collective effort to centralize and distribute phenotype and genotype/genomics information (Fischbach & Lord, 2010). It is important to discuss some limitations of the present study. Although the emphasis was on selecting measures representing different features, not specific instruments or scales, we recognize that the selected input variables used in PCA analysis do not represent the unique/best selection or ASD-related phenotype’s full range. Other additional measures encompassing motor skills or language are being considered in follow-up work. The scales were used with normative scores for age, allowing the construction of a map of phenotypic heterogeneity, but studies with repeated measures over time could add information to the temporal stability and feature selection to map construction. The input data for PCA were collected using parent’s reports (ADI-R, CBCL and Vineland) and, therefore, are subject to bias. Another limitation is that the normative sample (SSC) consists of simplex families and does not necessarily represent all ASD genetic architectures.

This work offers a series of contributions to research and practice. First, from the research point of view, we propose a model to observe the severity of a patient considering the heterogeneity of clinical phenotypic relationships, and provide patient placement in an ASD severity spectrum, allowing reliable case comparisons even among time-points of the same individual. Moreover, the approach proposed here presents an important differential: the idea of looking at a single subject. This can be important for treatment proposals and evaluation since individuals with ASD often differ in response to treatment. These findings may facilitate the development of more effective therapeutic strategies to optimize long-term results for individuals with ASD in a personalized medicine view, as well as sample selection for clinical and biomarker studies.

## Supporting information

TEAplot tool

## Data Availability

Data that support the findings of this study are available by request to the Simons Foundation Autism Research Initiative (SFARI). Other data are not publicly available due to restrictions regarding the privacy of research participants.

## Acknowledgements

We would like to thank all the PROTEA (Autism Spectrum Disorders Program at the Psychiatric Institute of Hospital das Clínicas), TEAMM, and Mackenzie staff who worked on data collection as well as the participating families and patients. We would like to thank all our sources of funding. Part of this study was supported by the State of São Paulo Funding Agency (Fundação de Amparo à Pesquisa do Estado de São Paulo) under a special agreement with the Maria Cecília Souto Vidigal Foundation (grant number 2012/51584-0). Access to Simons Foundation Autism Research Initiative (SFARI) dataset was granted under the proper SFARI review process and also approved by Hospital das Clínicas da FMUSP Research Ethics Committee (CAPPesq) according to review number 2.647.063 and project CAAE57067016.2.0000.0068, review 1.637.312. The protocol was approved by the Research Ethics Committee of UNIFESP (REC) and registered at CAAE19927213.4.1001.5505. It was also financed in part by the Coordenação de Aperfeiçoamento de Pessoal de Nível Superior, Brazil (CAPES), Finance Code 001; and National Program to Support Health Care for Persons with Disabilities (PRONAS). H. B. and J.J.M. are National Research Council (CNPq) research fellows. S.C.C. has received consulting fees from Pfizer and honoraria for lectures from Lundbeck. The other authors declare no conflict of interest.

