## Supplementary Methods and Tables for "A normative model representing autistic individuals amidst Autism Spectrum Disorders phenotypic heterogeneity"

### SUPPLEMENTARY MATERIAL

### SUPPLEMENTARY METHODS

#### IQ quantification details for SSC

Concerning the SSC database, the variable Full-Scale IQ provides an estimate of the individual's overall cognitive ability and consists of both Deviation IQs (i.e. IQs derived from standard scores) and Ratio IQs (i.e. IQs derived by dividing their averaged mental ages by chronological age for individuals who were not able to obtain a Deviation IQ). If a participant had a Full-Scale Deviation IQ, this value was always prioritized. However, if a Full-Scale Deviation IQ was not available (e.g. the participant was not able to establish a basal on a cognitive test and was administered a test for which standard scores were not available for his/her age), the Full-Scale Ratio IQ was used.

For Brazilian patients' cognitive assessment the Total Intelligence Quotient (IQ) was selected (Laros et al. 2013 "Brazilian validation of the nonverbal intelligence test SON-R 2½-7[a]", Karino et al. 2011 "Evidences of convergent validity of SON-R 2½-7[a] with WPPSI-III and WISC-III") since it provides standardized and age-corrected estimates of overall cognitive ability validated in Portuguese language.

### Normative modeling

A 3-dimensional Gaussian distribution (also know as Multivariate Normal distribution) was fitted to the PCA-derived vector space. The actual data used to fit the model are not raw PCA results but rather a Z-score-like normalized vector space truncated (projected) at 3 dimensions. Estimates for PC1, PC2 and PC3 dimension's means and standard deviations were used to normalize the observation coordinates in terms of "standard deviations from the mean". This is the final TEAplot presentation in which the normative modeling is performed. Observed covariances among all three dimensions were approximately zero and forced as such. Therefore, the fitted model can be reduced to independent univariate N(0,1) normal distributions in each of the 3 principal component axes. The multivariate probability density function is spherically symmetrical.

The SSC (normative population) estimated parameters, 3 pairs of means and standard deviations, are used to transform any new observation from the original PCA-rotated space to the TEAplot standardized space.

Due to its spherical symmetry, the 3-dimensional probability density function can be easily rotated in any direction, in particular, the defined severity axis which goes from the all negative octant to the all positive octant (FigureS10). Therefore we can calculate the theoretical quantile of a given point in space simply using a straightforward N(0,1) integral:

$$M = \int_{-\infty}^{q} \int_{-\infty}^{\infty} \int_{-\infty}^{\infty} (2\pi)^{\frac{-3}{2}} \, e^{\frac{-1}{2}(r_1^2 + r_2^2 + r_3^2)} \, \mathrm{d}r_1 \, \mathrm{d}r_2 \, \mathrm{d}r_3 \qquad \text{eq. 1}$$

$$M = \int_{-\infty}^{q} \frac{1}{\sqrt{2\pi}} e^{\frac{-1}{2}r^2} \, dr \qquad \text{eq. 2}$$

where $r_1$, $r_2$ and $r_3$ are the coordinates of a patient in a rotated 3-dimensional coordinate system that aligns $r_3 = r$ along the severity axis, which in turn follows the main diagonal of direction (1,1,1) in the TEAplot. Using this symmetry argument, the value $q$ is just the distance between the plane $Ax_1+Bx_2+Cx_3+D = 0$ orthogonal to the severity axis which contains the 3-dimensional point representing the patient:

$$q = \frac{D}{\sqrt{A^2 + B^2 + C^2}} \qquad \text{eq. 3}$$

where $x_1$, $x_2$ and $x_3$ are the new patient coordinates in TEAplot (thus Z-score normalized PCA space 3-dimensional projection).

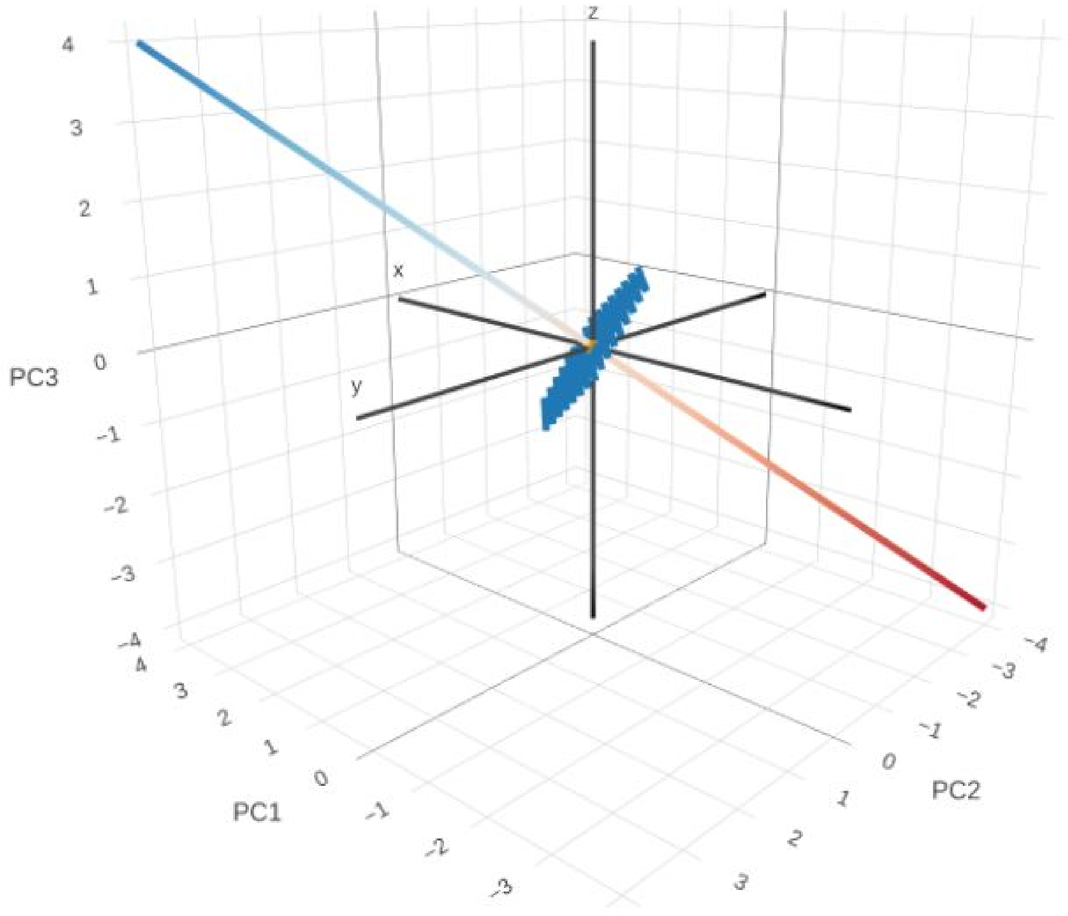

**Illustration 1 -** A$x_1$+B$x_2$+C$x_3$+D = 0 plane (blue) normal to the severity axis (diagonal red-to-blue line) is shown in TEAplot. In this specific example, the origin (0,0,0) lies in the plane and, therefore: M = 0.50 = 50%, MSS = 5.0 and $q$ = 0, since it splits the spherically symmetric multivariate normal probability density function in exactly half.

The equation 2 returns the cumulative probability of all observations up to the new observation TEAplot point for which we want to determine a deviation index concerning the reference sample (MSS):

$$MSS = 10\,M \qquad \text{eq. 4}$$

A patient may present some level of imbalance among the functional-cognitive-behavioral variation axes, presenting more skills in certain domains than in others. Therefore, to assign an imbalance score of phenotypic presentation, we created an index, Imbalance Score (ImS), based on a straightforward entropy calculation:

$$ImS = -10\,p_1 \log_3(p_1) - 10\,p_2 \log_3(p_2) - 10\,p_3 \log_3(p_3) \qquad \text{eq. 5}$$

where: $p_1 = |x_1|/(|x_1|+|x_2|+|x_3|)$, $p_2 = |x_2|/(|x_1|+|x_2|+|x_3|)$ and $p_3 = |x_3|/(|x_1|+|x_2|+|x_3|)$, and $x_1$, $x_2$ and $x_3$ are the patient coordinates in TEAplot.

**SUPPLEMENTARY TABLES**

**TableS1: Means and standard deviations from the scores obtained by patients with ASD from the SSC sample in the tests, scales, and questionnaires used as input for the PCA.**

| | | |
|---|---|---|
| ADI – R | | |
| | Socialization | 20.32 (5.70) |
| | Communication | 15.95 (4.31) |
| | Restricted and repetitive behavior | 6.52 (2.49) |
| Full-scale IQ | | |
| | IQ | 81.23 (27.9) |
| VABS | | |
| | Communication | 77.12 (14.49) |
| | Socialization | 70.98 (12.55) |
| | Daily Living Skills | 76.45 (13.83) |
| CBCL Problems índex* | | |
| | Internalizing Problems | 60.31 (9.55) |
| | Externalizing Problems | 56.57 (10.6) |
| *Data are T-scores | | |

Note: VABS = Vineland Adaptive Behavior Scales; ADI-R = Autism Diagnostic Interview-Revised; IQ = Intelligence Quotient; CBCL = Child Behavior Checklist.

**Table S2: Variance explained by each principal component when using the sub-items of VABS, ADI-R, total IQ and internalizing and externalizing CBCL evaluated in 2744 SSC patients as input measures.**

| **Components** | **1** | **2** | **3** | **4** | **5** | **6** | **7** | **8** | **9** |
|---|---|---|---|---|---|---|---|---|---|
| Eigenvalue | 3.49 | 1.65 | 1.39 | 0.74 | 0.5 | 0.43 | 0.38 | 0.23 | 0.19 |
| Explained Percentage of Variance | 38.7 | 18.3 | 15.4 | 8.2 | 5.6 | 4.7 | 4.2 | 2.6 | 2.2 |
| Cumulative Explained Percentage of Variance | 38.7 | 57.0 | 72.5 | 80.7 | 86.2 | 91.0 | 95.2 | 97.8 | 100 |
| **Correlations** | | | | | | | | | |
| VABS Communication | 0.89 | -0.16 | 0.17 | -0.03 | 0.06 | 0.0 | -0.09 | 0.16 | -0.34 |
| VABS Daily Living Skills | 0.85 | -0.13 | 0.19 | -0.05 | 0.0 | 0.31 | -0.05 | -0.35 | 0.02 |
| VABS Socialization | 0.87 | 0.04 | 0.12 | 0.05 | 0.15 | 0.2 | 0.27 | 0.24 | 0.2 |
| ADI-R Socialization | -0.69 | -0.18 | 0.43 | -0.28 | -0.11 | 0.37 | -0.24 | 0.15 | 0.03 |
| ADI-R Communication | -0.31 | -0.34 | 0.73 | -0.32 | 0.21 | -0.21 | 0.24 | -0.08 | -0.02 |
| ADI-R Restricted and Repetitive Behavior | -0.17 | -0.52 | 0.45 | 0.7 | -0.07 | -0.02 | -0.04 | 0.01 | 0.02 |
| CBCL Externalizing Problems | -0.14 | -0.71 | -0.48 | -0.03 | 0.47 | 0.03 | -0.16 | 0 | 0.04 |

| | | | | | | | | | |
|---|---|---|---|---|---|---|---|---|---|
| CBCL Internalizing Problems | -0.04 | -0.78 | -0.39 | -0.15 | -0.38 | 0.08 | 0.26 | 0.01 | -0.04 |
| Total IQ | 0.78 | -0.29 | 0.12 | -0.2 | -0.2 | -0.32 | -0.29 | 0.04 | 0.18 |

Note: VABS = Vineland Adaptive Behavior Scales; ADI-R = Autism Diagnostic Interview-Revised; IQ = Intelligence Quotient; CBCL = Child Behavior Checklist.

**Table S3: PCA explained variances and loadings when using 823 SSC patients**. *Similar to Table S2 however randomly sampling 30% of SSC patients.*

| **Components** | **1** | **2** | **3** | **4** | **5** | **6** | **7** | **8** | **9** |
|---|---|---|---|---|---|---|---|---|---|
| Eigenvalue | 3.53 | 1.74 | 1.34 | 0.76 | 0.48 | 0.37 | 0.37 | 0.22 | 0.19 |
| Explained Percentage of Variance | 39.2 | 19.32 | 14.85 | 8.40 | 5.36 | 4.15 | 4.10 | 2.46 | 2.15 |
| Cumulative Explained Percentage of Variance | 39.2 | 58.53 | 73.38 | 81.8 | 87.14 | 91.29 | 95.4 | 97.8 | 100 |
| **Correlations** | | | | | | | | | |
| VABS Communication | 0.89 | -0.21 | 0.15 | -0.03 | 0.05 | -0.04 | -0.02 | 0.18 | -0.33 |
| VABS Daily Living Skills | 0.87 | -0.15 | 0.16 | -0.04 | -0.06 | -0.23 | 0.09 | -0.36 | -0.01 |
| VABS Socialization | 0.88 | 0.02 | 0.10 | 0.04 | 0.12 | -0.03 | 0.35 | 0.19 | 0.22 |
| ADI-R Socialization | -0.69 | -0.27 | 0.41 | -0.27 | -0.15 | -0.43 | 0.02 | 0.13 | 0.03 |
| ADI-R Communication | -0.31 | -0.43 | 0.69 | -0.30 | 0.24 | 0.29 | 0.09 | -0.08 | -0.01 |
| ADI-R Restricted and Repetitive Behavior | -0.13 | -0.57 | 0.37 | 0.72 | -0.09 | 0.01 | -0.05 | 0.01 | 0.03 |
| CBCL Externalizing Problems | -0.16 | -0.66 | -0.54 | 0.00 | 0.47 | -0.15 | -0.05 | -0.01 | 0.02 |
| CBCL Internalizing Problems | -0.08 | -0.73 | -0.46 | -0.16 | -0.39 | 0.15 | 0.21 | 0.00 | -0.02 |
| Total IQ | 0.78 | -0.33 | 0.07 | -0.22 | -0.11 | 0.06 | -0.42 | 0.06 | 0.18 |

Note: VABS = Vineland Adaptive Behavior Scales; ADI-R = Autism Diagnostic Interview-Revised; IQ = Intelligence Quotient; CBCL = Child Behavior Checklist.

**Table S4: PCA explained variances and loadings when using 1098 SSC patients.** *Similar to Table S2 however randomly sampling 40% of SSC patients.*

| **Components** | **1** | **2** | **3** | **4** | **5** | **6** | **7** | **8** | **9** |
|---|---|---|---|---|---|---|---|---|---|
| Eigenvalue | 3.45 | 1.68 | 1.38 | 0.72 | 0.50 | 0.46 | 0.38 | 0.24 | 0.20 |
| Explained Percentage of Variance | 38.35 | 18.63 | 15.36 | 7.97 | 5.51 | 5.11 | 4.22 | 2.64 | 2.21 |
| Cumulative Explained Percentage of Variance | 38.35 | 56.98 | 72.34 | 80.30 | 85.82 | 90.92 | 95.15 | 97.79 | 100 |
| **Correlations** | | | | | | | | | |
| VABS Communication | -0.88 | 0.18 | -0.17 | 0.03 | -0.05 | -0.02 | 0.09 | -0.20 | 0.33 |
| VABS Daily Living Skills | -0.84 | 0.16 | -0.21 | 0.00 | -0.03 | 0.30 | 0.14 | 0.35 | 0.01 |
| VABS Socialization | -0.87 | -0.03 | -0.12 | -0.04 | -0.19 | 0.24 | -0.19 | -0.22 | -0.23 |
| ADI-R Socialization | 0.70 | 0.22 | -0.42 | 0.21 | 0.11 | 0.30 | 0.34 | -0.15 | -0.05 |
| ADI-R Communication | 0.33 | 0.36 | -0.70 | 0.38 | -0.18 | -0.12 | -0.29 | 0.07 | 0.03 |
| ADI-R Restricted and Repetitive Behavior | 0.20 | 0.55 | -0.41 | -0.69 | -0.02 | -0.09 | 0.01 | -0.01 | -0.03 |
| CBCL Externalizing Problems | 0.11 | 0.68 | 0.52 | 0.13 | -0.44 | -0.08 | 0.17 | 0.00 | -0.04 |
| CBCL Internalizing Problems | 0.03 | 0.77 | 0.41 | 0.07 | 0.36 | 0.20 | -0.24 | -0.02 | 0.04 |
| Total IQ | -0.76 | 0.28 | -0.14 | 0.19 | 0.29 | -0.39 | 0.17 | -0.01 | -0.17 |

Note: VABS = Vineland Adaptive Behavior Scales; ADI-R = Autism Diagnostic Interview-Revised; IQ = Intelligence Quotient; CBCL = Child Behavior Checklist.

**Table S5: PCA explained variances and loadings when using 1372 SSC patients.** *Similar to Table S2 however randomly sampling 50% of SSC patients.*

| **Components** | **1** | **2** | **3** | **4** | **5** | **6** | **7** | **8** | **9** |
|---|---|---|---|---|---|---|---|---|---|
| Eigenvalue | 3.46 | 1.65 | 1.38 | 0.75 | 0.50 | 0.43 | 0.38 | 0.25 | 0.20 |
| Explained Percentage of Variance | 38.48 | 18.33 | 15.39 | 8.37 | 5.51 | 4.74 | 4.25 | 2.73 | 2.21 |
| Cumulative Explained Percentage of Variance | 38.48 | 56.80 | 72.19 | 80.56 | 86.08 | 90.81 | 95.07 | 97.79 | 100 |
| **Correlations** | | | | | | | | | |
| VABS Communication | -0.88 | 0.17 | -0.18 | 0.02 | -0.06 | 0.04 | -0.09 | 0.25 | 0.30 |
| VABS Daily Living Skills | -0.85 | 0.14 | -0.19 | 0.04 | -0.01 | -0.28 | -0.15 | -0.35 | 0.06 |
| VABS Socialization | -0.86 | -0.03 | -0.12 | -0.04 | -0.20 | -0.27 | 0.17 | 0.18 | -0.25 |
| ADI-R Socialization | 0.69 | 0.15 | -0.44 | 0.27 | 0.15 | -0.28 | -0.33 | 0.15 | -0.06 |
| ADI-R Communication | 0.32 | 0.29 | -0.74 | 0.33 | -0.25 | 0.14 | 0.26 | -0.08 | 0.03 |
| ADI-R Restricted and Repetitive Behavior | 0.22 | 0.46 | -0.47 | -0.72 | 0.05 | 0.02 | -0.02 | 0.00 | -0.02 |
| CBCL Externalizing Problems | 0.16 | 0.73 | 0.46 | 0.04 | -0.42 | 0.07 | -0.22 | 0.00 | -0.06 |
| CBCL Internalizing Problems | 0.10 | 0.81 | 0.33 | 0.12 | 0.30 | -0.19 | 0.28 | 0.02 | 0.05 |
| Total IQ | -0.75 | 0.32 | -0.15 | 0.18 | 0.30 | 0.37 | -0.13 | 0.00 | -0.18 |

Note: VABS = Vineland Adaptive Behavior Scales; ADI-R = Autism Diagnostic Interview-Revised; IQ = Intelligence Quotient; CBCL = Child Behavior Checklist.

**Table S6: PCA explained variances and loadings when using 1646 SSC patients.** *Similar to Table S2 however randomly sampling 60% of SSC patients.*

| **Components** | **1** | **2** | **3** | **4** | **5** | **6** | **7** | **8** | **9** |
|---|---|---|---|---|---|---|---|---|---|
| Eigenvalue | 3.47 | 1.65 | 1.37 | 0.75 | 0.50 | 0.44 | 0.39 | 0.24 | 0.19 |
| Explained Percentage of Variance | 38.56 | 18.36 | 15.18 | 8.36 | 5.60 | 4.87 | 4.30 | 2.66 | 2.11 |
| Cumulative Explained Percentage of Variance | 38.56 | 56.92 | 72.10 | 80.47 | 86.06 | 90.93 | 95.23 | 97.89 | 100 |
| **Correlations** | | | | | | | | | |
| VABS Communication | 0.89 | -0.15 | 0.16 | -0.04 | 0.07 | 0.00 | 0.10 | 0.19 | 0.33 |
| VABS Daily Living Skills | 0.85 | -0.11 | 0.20 | -0.06 | -0.02 | 0.29 | 0.08 | -0.36 | 0.00 |
| VABS Socialization | 0.86 | 0.03 | 0.12 | 0.04 | 0.14 | 0.26 | -0.25 | 0.22 | -0.21 |
| ADI-R Socialization | -0.68 | -0.18 | 0.43 | -0.29 | -0.15 | 0.34 | 0.28 | 0.14 | -0.04 |
| ADI-R Communication | -0.30 | -0.33 | 0.74 | -0.31 | 0.25 | -0.19 | -0.25 | -0.07 | 0.02 |
| ADI-R Restricted and Repetitive Behavior | -0.13 | -0.53 | 0.44 | 0.71 | -0.09 | -0.01 | 0.03 | 0.01 | -0.02 |
| CBCL Externalizing Problems | -0.13 | -0.72 | -0.47 | -0.02 | 0.46 | 0.04 | 0.18 | 0.00 | -0.05 |
| CBCL Internalizing Problems | -0.02 | -0.78 | -0.37 | -0.18 | -0.37 | 0.07 | -0.27 | 0.00 | 0.05 |
| Total IQ | 0.78 | -0.26 | 0.12 | -0.19 | -0.20 | -0.37 | 0.25 | 0.04 | -0.18 |

Note: VABS = Vineland Adaptive Behavior Scales; ADI-R = Autism Diagnostic Interview-Revised; IQ = Intelligence Quotient; CBCL = Child Behavior Checklist.

**Table S7: PCA explained variances and loadings when using 1921 SSC patients.** *Similar to Table S2 however randomly sampling 70% of SSC patients.*

| **Components** | **1** | **2** | **3** | **4** | **5** | **6** | **7** | **8** | **9** |
|---|---|---|---|---|---|---|---|---|---|
| Eigenvalue | 3.55 | 1.63 | 1.35 | 0.76 | 0.51 | 0.41 | 0.38 | 0.22 | 0.19 |
| Explained Percentage of Variance | 39.34 | 18.1 | 15.02 | 8.47 | 5.63 | 4.57 | 4.22 | 2.49 | 2.12 |
| Cumulative Explained Percentage of Variance | 39.34 | 57.5 | 72.50 | 80.97 | 86.60 | 91.16 | 95.39 | 97.88 | 100 |
| **Correlations** | | | | | | | | | |
| VABS Communication | 0.89 | -0.16 | 0.19 | -0.02 | 0.05 | 0.00 | -0.09 | -0.13 | 0.35 |
| VABS Daily Living Skills | 0.86 | -0.13 | 0.18 | -0.05 | 0.04 | 0.30 | -0.01 | 0.34 | -0.03 |
| VABS Socialization | 0.87 | 0.04 | 0.12 | 0.05 | 0.18 | 0.16 | 0.28 | -0.25 | -0.18 |
| ADI-R Socialization | -0.70 | -0.16 | 0.40 | -0.31 | -0.06 | 0.40 | -0.20 | -0.14 | -0.02 |
| ADI-R Communication | -0.34 | -0.30 | 0.74 | -0.30 | 0.19 | -0.25 | 0.23 | 0.08 | 0.01 |
| ADI-R Restricted and Repetitive Behavior | -0.20 | -0.48 | 0.47 | 0.71 | -0.11 | 0.02 | -0.04 | -0.01 | -0.03 |
| CBCL Externalizing Problems | -0.14 | -0.74 | -0.42 | 0.01 | 0.48 | 0.00 | -0.16 | 0.00 | -0.04 |
| CBCL Internalizing Problems | -0.04 | -0.78 | -0.37 | -0.16 | -0.37 | 0.06 | 0.28 | -0.01 | 0.04 |
| Total IQ | 0.77 | -0.29 | 0.16 | -0.21 | -0.23 | -0.25 | -0.32 | -0.05 | -0.18 |

Note: VABS = Vineland Adaptive Behavior Scales; ADI-R = Autism Diagnostic Interview-Revised; IQ = Intelligence Quotient; CBCL = Child Behavior Checklist.

**Table S8: PCA explained variances and loadings when using 2195 SSC patients.** *Similar to Table S2 however randomly sampling 80% of SSC patients.*

| **Components** | **1** | **2** | **3** | **4** | **5** | **6** | **7** | **8** | **9** |
|---|---|---|---|---|---|---|---|---|---|
| Eigenvalue | 3.45 | 1.68 | 1.40 | 0.73 | 0.50 | 0.43 | 0.38 | 0.23 | 0.20 |
| Explained Percentage of Variance | 38.28 | 18.63 | 15.59 | 8.10 | 5.59 | 4.74 | 4.25 | 2.60 | 2.21 |
| Cumulative Explained Percentage of Variance | 38.28 | 56.91 | 72.51 | 80.61 | 86.20 | 90.94 | 95.19 | 97.79 | 100 |
| **Correlations** | | | | | | | | | |
| VABS Communication | 0.89 | -0.18 | 0.15 | -0.04 | 0.06 | -0.01 | -0.10 | -0.13 | 0.35 |
| VABS Daily Living Skills | 0.85 | -0.13 | 0.19 | -0.04 | 0.00 | -0.30 | -0.03 | 0.35 | -0.04 |
| VABS-Socialization | 0.87 | 0.04 | 0.12 | 0.08 | 0.14 | -0.20 | 0.26 | -0.25 | -0.19 |
| ADI-R Socialization | -0.67 | -0.23 | 0.42 | -0.28 | -0.11 | -0.38 | -0.24 | -0.14 | -0.03 |
| ADI-R Communication | -0.30 | -0.42 | 0.69 | -0.30 | 0.25 | 0.21 | 0.25 | 0.07 | 0.01 |
| ADI-R Restricted and Repetitive Behavior | -0.15 | -0.57 | 0.41 | 0.69 | -0.12 | 0.03 | -0.05 | -0.01 | -0.02 |
| CBCL Externalising problems | -0.11 | -0.67 | -0.54 | 0.01 | 0.47 | -0.05 | -0.17 | 0.00 | -0.05 |
| CBCL Internalising Problems | -0.02 | -0.74 | -0.46 | -0.18 | -0.37 | -0.06 | 0.27 | -0.01 | 0.04 |
| Total IQ | 0.77 | -0.29 | 0.11 | -0.23 | -0.19 | 0.32 | -0.28 | -0.05 | -0.18 |

Note: VABS = Vineland Adaptive Behavior Scales; ADI-R = Autism Diagnostic Interview-Revised; IQ = Intelligence Quotient; CBCL = Child Behavior Checklist.

**Table S9. Means and standard deviations of the scores obtained by the case presentation patients in the tests, scales and questionnaires.**

| | | |
|---|---|---|
| ADI – R | | |
| | Socialization | 23.87 (3.92) |
| | Communication | 12.98 (3.36) |
| | Restricted repetitive behavior | 5.91 (2.64) |
| SON-R | | |
| | IQ | 58.5 (8.81) |
| VABS | | |
| | Communication | 47.06( 9.02) |
| | Socialization | 54.23 (10.87) |
| | Daily Living Skills | 50.41 (13.28) |
| CBCL Problems índex* | | |
| | Internalizing Problems | 67 (8.65) |
| | Externalizing Problems | 64.35 (9.12) |
| *Data are T-scores | | |

Note: VABS = Vineland Adaptive Behavior Scales; ADI-R = Autism Diagnostic Interview-Revised; IQ = Intelligence Quotient; CBCL = Child Behavior Checklist; SON-R: Snijders-Oomen nonverbal intelligence tests.

**Table S10: Case presentation: TEAplot coordinates, severity and imbalance scores**

| IDs | VABS Com | VABS Soc | VABS DLS | ADI Soc | ADI Com | ADI RRB | Total IQ | CBCL Int | CBCL Ext | PC1 | PC2 | PC3 | MSS | ImS |
|---|---|---|---|---|---|---|---|---|---|---|---|---|---|---|
| ID - 01 | 39 | 48 | 37 | 30 | 14 | 13 | 49 | 76 | 67 | -2.55 | -1.46 | 0.65 | 0.3 | 8.8 |
| ID - 02 | 39 | 32 | 88 | 27 | 14 | 10 | 50 | 80 | 77 | -1.84 | -1.94 | 1.25 | 0.7 | 9.8 |
| ID - 04 | 34 | 48 | 32 | 27 | 16 | 5 | 49 | 74 | 57 | -2.47 | 0.07 | 1.31 | 2.6 | 6.6 |
| ID - 05 | 54 | 51 | 53 | 25 | 13 | 8 | 66 | 64 | 60 | -1.48 | -0.10 | 0.74 | 3.1 | 7.2 |
| ID - 07 | 67 | 59 | 56 | 24 | 13 | 10 | 55 | 79 | 54 | -1.13 | -0.86 | 0.62 | 2.1 | 9.7 |
| ID - 08 | 52 | 54 | 59 | 25 | 13 | 10 | 67 | 58 | 56 | -1.36 | 0.07 | 0.11 | 2.5 | 4.2 |
| ID - 09 | 49 | 61 | 54 | 21 | 5 | 6 | 71 | 59 | 56 | -0.95 | 1.03 | 1.86 | 8.7 | 9.6 |
| ID - 10 | 50 | 51 | 55 | 21 | 14 | 4 | 72 | 65 | 63 | -1.28 | 0.23 | 1.48 | 6.0 | 8.3 |
| ID - 11 | 51 | 54 | 63 | 23 | 20 | 5 | 72 | 61 | 63 | -1.27 | -0.06 | 0.29 | 2.7 | 5.8 |
| ID - 12 | 39 | 42 | 51 | 26 | 12 | 4 | 51 | 66 | 67 | -2.04 | 0.23 | 1.87 | 5.2 | 7.9 |
| ID - 13 | 52 | 66 | 60 | 18 | 8 | 7 | 73 | 76 | 77 | -0.75 | -0.92 | 2.58 | 7.0 | 8.6 |
| ID - 15 | 68 | 32 | 44 | 28 | 11 | 5 | 68 | 73 | 79 | -1.84 | -0.99 | 2.19 | 3.5 | 9.6 |
| ID - 16 | 49 | 54 | 56 | 24 | 13 | 10 | 55 | 74 | 71 | -1.60 | -1.18 | 1.22 | 1.8 | 9.9 |
| ID - 17 | 48 | 53 | 52 | 28 | 13 | 6 | 49 | 65 | 67 | -1.79 | -0.08 | 1.20 | 3.5 | 7.1 |
| ID - 19 | 37 | 45 | 21 | 30 | 14 | 8 | 56 | 66 | 69 | -2.77 | -0.36 | 1.23 | 1.4 | 7.8 |
| ID - 20 | 45 | 60 | 50 | 24 | 14 | 5 | 55 | 51 | 46 | -1.46 | 1.61 | 0.30 | 6.0 | 8.5 |
| ID - 21 | 51 | 55 | 62 | 24 | 13 | 10 | 55 | 61 | 52 | -1.36 | 0.17 | 0.13 | 2.7 | 5.6 |
| ID - 22 | 54 | 61 | 70 | 26 | 12 | 7 | 62 | 70 | 65 | -1.03 | -0.49 | 1.06 | 3.9 | 9.5 |
| ID - 23 | 63 | 68 | 84 | 24 | 13 | 10 | 51 | 58 | 54 | -0.53 | 0.08 | -0.27 | 3.4 | 8.2 |
| ID - 24 | 47 | 61 | 42 | 26 | 19 | 8 | 50 | 76 | 67 | -1.92 | -1.06 | 0.69 | 0.9 | 9.2 |
| ID - 25 | 40 | 65 | 54 | 28 | 14 | 3 | 58 | 80 | 77 | -1.60 | -0.88 | 2.18 | 4.3 | 9.5 |
| ID - 27 | 67 | 79 | 64 | 13 | 5 | 2 | 69 | 58 | 57 | 0.22 | 1.56 | 2.44 | 9.9 | 7.7 |
| ID - 28 | 48 | 53 | 59 | 14 | 10 | 5 | 69 | 73 | 70 | -0.96 | -0.22 | 2.65 | 8.0 | 7.0 |
| ID - 29 | 50 | 51 | 54 | 28 | 7 | 5 | 54 | 72 | 56 | -1.55 | 0.36 | 1.87 | 6.5 | 8.6 |

| | | | | | | | | | | | | | | |
|---|---|---|---|---|---|---|---|---|---|---|---|---|---|---|
| ID - 30 | 48 | 53 | 52 | 24 | 13 | 10 | 49 | 53 | 59 | -1.69 | 0.39 | 0.28 | 2.8 | 7.2 |
| ID - 31 | 33 | 49 | 31 | 25 | 14 | 9 | 49 | 72 | 67 | -2.49 | -0.59 | 1.42 | 1.7 | 8.7 |
| ID- 33 | 47 | 54 | 35 | 28 | 16 | 5 | 49 | 77 | 77 | -2.18 | -0.99 | 1.82 | 2.2 | 9.6 |
| ID - 34 | 45 | 61 | 67 | 21 | 17 | 4 | 49 | 73 | 76 | -1.26 | -0.69 | 1.71 | 4.4 | 9.5 |
| ID - 35 | 42 | 51 | 44 | 25 | 13 | 5 | 49 | 56 | 58 | -1.91 | 0.98 | 1.07 | 5.3 | 9.6 |
| ID - 36 | 50 | 63 | 52 | 27 | 12 | 4 | 55 | 72 | 65 | -1.42 | -0.06 | 1.67 | 5.4 | 7.0 |
| ID - 39 | 31 | 45 | 32 | 24 | 13 | 10 | 49 | 66 | 51 | -2.48 | 0.29 | 0.80 | 2.1 | 7.2 |
| ID - 42 | 50 | 54 | 52 | 20 | 12 | 6 | 50 | 62 | 57 | -1.39 | 0.64 | 1.31 | 6.3 | 9.6 |
| ID - 43 | 65 | 85 | 56 | 21 | 15 | 8 | 75 | 56 | 62 | -0.39 | 0.10 | 0.16 | 4.7 | 8.6 |
| ID - 44 | 49 | 74 | 53 | 13 | 13 | 3 | 52 | 53 | 60 | -0.70 | 1.46 | 1.64 | 9.2 | 9.5 |
| ID - 46 | 58 | 80 | 55 | 20 | 5 | 2 | 69 | 73 | 83 | -0.42 | -0.24 | 3.51 | 9.5 | 4.9 |
| ID - 47 | 49 | 61 | 49 | 24 | 20 | 8 | 66 | 60 | 64 | -1.56 | -0.31 | -0.02 | 1.4 | 4.6 |
| ID - 48 | 41 | 44 | 35 | 18 | 12 | 7 | 49 | 76 | 69 | -2.07 | -0.47 | 2.40 | 4.7 | 8.5 |
| ID - 49 | 51 | 53 | 55 | 20 | 14 | 9 | 71 | 70 | 64 | -1.31 | -0.66 | 1.04 | 3.0 | 9.7 |
| ID - 50 | 35 | 50 | 25 | 20 | 8 | 6 | 49 | 62 | 68 | -2.18 | 0.64 | 2.57 | 7.3 | 8.9 |
| ID - 51 | 48 | 56 | 45 | 27 | 13 | 3 | 54 | 45 | 56 | -1.66 | 1.75 | 0.73 | 6.8 | 9.4 |
| ID - 53 | 45 | 54 | 53 | 25 | 9 | 4 | 68 | 79 | 85 | -1.51 | -1.10 | 3.05 | 6.0 | 9.1 |
| ID - 54 | 52 | 56 | 50 | 23 | 12 | 3 | 75 | 71 | 66 | -1.24 | 0.008 | 2.00 | 6.7 | 6.2 |
| ID - 56 | 49 | 51 | 52 | 23 | 13 | 4 | 67 | 59 | 56 | -1.41 | 0.87 | 1.14 | 6.4 | 9.8 |
| ID - 57 | 38 | 37 | 53 | 28 | 14 | 2 | 55 | 70 | 55 | -2.12 | 0.60 | 1.52 | 5.0 | 9.0 |
| ID - 58 | 32 | 45 | 20 | 26 | 14 | 4 | 49 | 49 | 55 | -2.65 | 1.70 | 1.08 | 5.3 | 9.4 |
| ID - 59 | 49 | 52 | 47 | 26 | 13 | 8 | 56 | 74 | 62 | -1.79 | -0.56 | 1.17 | 2.5 | 9.1 |
| ID - 60 | 54 | 66 | 68 | 17 | 7 | 3 | 55 | 81 | 79 | -0.60 | -0.62 | 3.45 | 9.0 | 6.9 |
| ID - 61 | 47 | 52 | 52 | 23 | 11 | 5 | 60 | 59 | 66 | -1.50 | 0.50 | 1.61 | 6.4 | 9.1 |
| ID - 63 | 42 | 48 | 36 | 23 | 14 | 6 | 49 | 65 | 57 | -2.09 | 0.48 | 1.26 | 4.2 | 8.7 |
| ID - 65 | 28 | 21 | 46 | 28 | 15 | 7 | 54 | 81 | 65 | -2.91 | -0.93 | 1.67 | 1.0 | 9.1 |
| ID - 66 | 50 | 56 | 68 | 22 | 14 | 9 | 72 | 70 | 60 | -1.08 | -0.60 | 0.67 | 2.8 | 9.7 |

| | | | | | | | | | | | | | | |
|---|---|---|---|---|---|---|---|---|---|---|---|---|---|---|
| ID - 68 | 32 | 66 | 48 | 26 | 13 | 4 | 50 | 63 | 64 | -1.76 | 0.57 | 1.49 | 5.7 | 9.2 |
| ID - 69 | 34 | 48 | 23 | 30 | 13 | 2 | 49 | 69 | 61 | -2.62 | 0.67 | 1.97 | 5.1 | 8.9 |
| ID - 72 | 44 | 56 | 45 | 21 | 18 | 5 | 55 | 59 | 43 | -1.62 | 1.23 | 0.20 | 4.6 | 8.1 |
| ID - 73 | 61 | 53 | 62 | 22 | 14 | 3 | 60 | 61 | 65 | -1.04 | 0.42 | 1.36 | 6.7 | 9.1 |
| ID - 74 | 41 | 50 | 41 | 29 | 13 | 4 | 66 | 71 | 68 | -2.04 | -0.19 | 1.75 | 3.9 | 7.7 |
| ID - 75 | 46 | 52 | 55 | 28 | 14 | 4 | 62 | 78 | 77 | -1.72 | -1.01 | 2.00 | 3.4 | 9.7 |
| ID - 77 | 47 | 51 | 45 | 24 | 19 | 3 | 70 | 63 | 62 | -1.71 | 0.29 | 0.88 | 3.8 | 8.2 |
| ID - 78 | 47 | 57 | 53 | 23 | 20 | 6 | 59 | 65 | 67 | -1.60 | -0.38 | 0.56 | 2.1 | 8.3 |
| ID - 80 | 51 | 47 | 50 | 24 | 9 | 4 | 69 | 72 | 85 | -1.55 | -0.78 | 2.92 | 6.3 | 8.8 |

Note: VABS Com = Vineland Adaptive Behavior Scales Communication; VABS Soc = Vineland Adaptive Behavior Scales Socialization; VABS DLS = Vineland Adaptive Behavior Scales Daily Living Skills; ADI-R Soc = Autism Diagnostic Interview-Revised Socialization; ADI-R Com = Autism Diagnostic Interview-Revised Communication; ADI-R RRB = Autism Diagnostic Interview-Revised Repetitive Restricted Behavior; IQ = Intelligence Quotient; CBCL Int = Child Behavior Checklist Internalizing Problems; CBCL Ext = Child Behavior Checklist Externalizing Problems; PC = Principal Component (Z-score standardized); MSS = Multidimensional Severity Score; ImB = Imbalance Score

**Table S11: Coordinates of ID-28 trajectory: TEAplot, severity and imbalance scores.**

| Time | PC1 | PC2 | PC3 | MSS | Imbalance score |
|---|---|---|---|---|---|
| $t_1$ | -0.96 | -0.22 | 2.65 | 8.0 | 7.0 |
| $t_2$ | -0.82 | 2.15 | 1.02 | 9.1 | 9.2 |

Note: PC = Principal Component; MSS = Multidimensional Severity Score.

### SUPPLEMENTARY FIGURES

**Figure S1: Relationship between Vineland Adaptive Behavior Scales (VABS) Communication and the first three principal components coordinate system.**

**Figure S2: Relationship between Vineland Adaptive Behavior Scales (VABS) Socialization and the first three principal components coordinate system**

**Figure S3: Relationship between Vineland Adaptive Behavior Scales (VABS) Daily Living Skills and the first three principal components coordinate system**

**Figure S4: Relationship between Total Intelligence Quotient (IQ) and the first three principal components coordinate system**

**Figure S5: Relationship between Autism Diagnostic Interview-Revised (ADI-R) Socialization and the first three principal components coordinate system**

**Figure S6: Relationship between Child Behavior Checklist (CBCL) Internalizing Problems and the first three principal components coordinate system**

**Figure S7: Relationship between Child Behavior Checklist (CBCL) Externalizing Problems and the first three principal components coordinate system**

**Figure S8: Relationship between Autism Diagnostic Interview-Revised (ADI-R) Restricted and Repetitive Behavior  and the first three principal components coordinate system**

**Figure S9: Relationship between Autism Diagnostic Interview-Revised (ADI-R) Communication and the first three principal components coordinate system**

**Figure S10: Axis of phenotypic variation that represents the severity of the clinical presentation**. The line is directed from worst (all negative, red) to better (all positive, blue) and crosses the origin.

**Figure S11: Relationship between Vineland Adaptive Behavior Scales (VABS) Communication and the first three principal components coordinate system, normalized to Z-Scores.** The line is directed from worst (all negative, red) to better (all positive, blue) and crosses the origin.

**Figure S12: Relationship between Vineland Adaptive Behavior Scales (VABS) Socialization and the first three principal components coordinate system, normalized to Z-Scores.** The line is directed from worst (all negative, red) to better (all positive, blue) and crosses the origin.

**Figure S13: Relationship between Vineland Adaptive Behavior Scales (VABS) Daily Living Skills and the first three principal components coordinate system, normalized to Z-Scores.** The line is directed from worst (all negative, red) to better (all positive, blue) and crosses the origin.

**Figure S14: Relationship between Total Intelligence Quotient (IQ) and the first three principal components coordinate system, normalized to Z-Scores.** The line is directed from worst (all negative, red) to better (all positive, blue) and crosses the origin.

**Figure S15: Relationship between Autism Diagnostic Interview-Revised (ADI-R) Socialization and the first three principal components coordinate system, normalized to Z-Scores.** The line is directed from worst (all negative, red) to better (all positive, blue) and crosses the origin.

**Figure S16: Relationship between Child Behavior Checklist (CBCL) Internalizing Problems and the first three principal components coordinate system, normalized to Z-Scores.** The line is directed from worst (all negative, red) to better (all positive, blue) and crosses the origin.

**Figure S17: Relationship between Child Behavior Checklist (CBCL) Externalizing Problems and the first three principal components coordinate system, normalized to Z-Scores.** The line is directed from worst (all negative, red) to better (all positive, blue) and crosses the origin.

**Figure S18: Relationship between Autism Diagnostic Interview-Revised (ADI-R) Restricted and Repetitive Behavior and the first three principal components coordinate system, normalized to Z-Scores.** The line is directed from worst (all negative, red) to better (all positive, blue) and crosses the origin.

**Figure S19: Relationship between Autism Diagnostic Interview-Revised (ADI-R) Communication and the first three principal components coordinate system, normalized to Z-Scores.** The line is directed from worst (all negative, red) to better (all positive, blue) and crosses the origin.

**Figure S20: Histogram of the Multidimensional Severity Scores (MSS) values in the SSC sample.**

**Figure S21: Relationship between the Multidimensional Severity Scores (MSS) and ADOS-2 Calibrated Severity Scores (CSS).**

**Figure S22: Visualization of patient ID-28 on the map of phenotypic heterogeneity at two different moments in time.**
